# The Impact of Touch Interventions on Brain Activity in Moderately Preterm Infants: study protocol for a pilot randomized controlled trial

**DOI:** 10.1101/2025.03.30.25324897

**Authors:** Andrea Manzotti, Francesco Cerritelli, Erica Lombardi, Tansini Laura, Desdemona Pisanu, Debora Di Leo, Elisa Vergani, Andrea Righini, Filippo Arrigoni, Vassilios Fanos, Maria Rescigno, Pierangelo Veggiotti, Gianluca Lista, Diego Gazzolo

## Abstract

**Introduction:** Improving the quality of life for preterm children is a global health priority, given their vulnerability to neurocognitive impairments and adverse health consequences. Lack of post-hospital care further exacerbates these risks, necessitating effective interventions during the neonatal period. This protocol for a pilot study aims to investigate the effects of touch interventions, including physiotherapy and manual therapy (OMT), on brain activity in moderately preterm infants using brain functional magnetic resonance imaging (fMRI), computerized electroencephalogram (EEG), and metabolomics.

**Methods and analysis:** A 3-arm randomized sham-controlled trial will be conducted with 15 infants per experimental group. The study will include stable preterm infants born between 32.0 and 33.6 weeks of gestational age (GA) who do not require other intensive care treatments. Exclusion criteria encompass preterm infants born before 32.0 weeks GA, those born after 33.6 weeks GA, and newborns with respiratory and neurological pathologies.

The study aims to assess the impact of touch interventions on brain activity and metabolic sequelae. Using fMRI will primarily examine the pre-post changes in functional connectivity of the brain, while EEG will secondarily explore the preterm brain’s neural effects on slow delta waves band. Metabolomics will provide data on the effects among the 3 groups on metabolic changes associated with touch interventions.

**Ethics and dissemination:** Ethical approval has been obtained from the Ethics Committee of the local health agency in Milan (CET 449-2024). Understanding the effects of touch interventions on brain activity in moderately preterm infants, without needs of intensive care, can contribute to improving their clinical outcomes and promoting their growth, development, and social behavior. Findings from this pilot study will pave the way for future research, enabling the development of evidence-based interventions to enhance preterm infants’ well-being and long-term outcomes.

**Trial registration:** The research protocol has been registered on ClinicalTrials.gov (NCT05853991)

**Article Summary:** Strengths and limitations of this study

- The study presents rigorous study design with blinding measures to determine the effect of therapeutic touch as compared to affective and static touch
- The study employs a combination of high-resolution 3T fMRI, electrophysiology, and metabolomic analysis, providing objective and quantifiable biomarkers of neural and metabolomic responses to touch interventions.
- The study recruits a small sample size, which therefore might limited the generalizability of results
- The study assesses immediate post-intervention effects but does not include long-term neurodevelopmental follow-up

## INTRODUCTION

Complementary treatments, particularly touch-based therapies, are considered beneficial and ecologically sound in pediatrics [1]. Studies have shown that a specific touch modality known as “gentle touch” can decrease stress levels in premature infants [2,3]. This gentle touch has direct effects on the autonomic nervous system (ANS) of the child, reducing cortisol levels and potentially improving adaptability and health in preterm infants [4–7]. Gentle touch stimulates the skin’s C-tactile (CT) fibers, which are part of the interoceptive system responsible for collecting afferent signals from the body and integrating them in the insular cortex [1,7,8].

Touch-based therapies, such as osteopathic manipulative treatment (OMT), aim to improve the overall well-being of children who experience extended periods of immobility and reduced sensory stimulation. These therapies promote bodily self-regulation through manual techniques that correct somatic dysfunctions [9– 11]. OMT include joint mobilization, myofascial procedures, balanced ligament tensions, and cranial osteopathic techniques. OMT has been studied in various pediatric clinical conditions, including asthma, attention deficit hyperactivity disorder (ADHD), chronic constipation, scoliosis, otitis media, and autism, showing positive results [12–17]. In neonatology, OMT has been found to reduce hospitalization length and related costs and improve pain management in preterm infants [6,18,19].

Early intervention with OMT has been shown to provide greater benefits for newborns. Lanaro et al. (2017) found that premature babies with a gestational age of less than 32 weeks experienced a significant reduction in hospital stay length after manual intervention, with a reduction of nearly nine days[6]. However, the neurophysiological and neurobiological mechanisms underlying these clinical improvements are not well understood, as neuroimaging studies have been limited, primarily focusing on the adult population.

Studies have indicated that OMT may lead to changes in brain perfusion and functional connectivity. Shi and colleagues (2011) observed a progressive decrease in oxygen saturation in the prefrontal lobes bilaterally after OMT[20]. Cerritelli and colleagues (2017 and 2020) demonstrated significant effects on functional connectivity patterns in cortical areas involved in processing touch-related interoceptive and attentional value[21,22]. In another trial, Cerritelli and coauthors (2021) found that OMT had both central and peripheral effects, including changes in cerebral blood flow in areas associated to pain processing and modulation and heart rate variability[23]. Results confirmed by a recent study, where OMT was associated with a reduction in connectivity within a parietal cluster encompassing the somatosensory cortex, alongside enhanced connectivity in the right anterior insula and both ventral and dorsal anterolateral prefrontal cortices. Importantly, authors found that the increase in connectivity strength within the ventral anterolateral prefrontal cortex was significantly correlated with the observed reduction in pain perception following OMT[24]. While previous research mainly focused on chronic effects of OMT, Tamburella et al. (2019) and Tramontano et al. (2020) explored the acute effects of OMT on healthy subjects[25,26]. They observed changes in blood flow and reported larger cerebral blood flow changes with extended osteopathic treatment, along with associated changes in the autonomic response. Despite these findings, the effects of OMT on brain activity in newborns remain unexplored.

Neuroimaging techniques are crucial for understanding brain development and plasticity. Electroencephalography (EEG) is a reliable method to study brain function, neural plasticity, and metabolic phenomena [27,28]. Although EEG has long been used for neurodevelopmental assessment and prognosis, its potential for therapeutic monitoring has been underutilized [29]. EEG allows the exploration of brain activity during the preterm period, characterized by transient spontaneous activities (SAT) and slow delta waves (F 0.3-1.5 Hz) with superimposed fast alpha and beta activities [30–32]. The low spectral power of delta waves in preterm infants may indicate impaired brain development due to premature exposure to the extrauterine environment [30,33]. However, external somatosensory stimuli, such as gentle touch, can stimulate delta waves and modify brain electrical activity, particularly in perirolandic regions [34–36].

Recent studies have reported changes in brain maturation in preterm infants undergoing touch-based interventions, showing increased EEG spectral power and delta activity in frontopolar regions [37–39]. Infant massage has also been shown to promote premature brain maturation by stimulating electrical activity and increasing the amplitude of delta waves [40]. Touch interventions have a topographic distribution in terms of maturation changes, affecting regions, such as somatosensory cortex and insula, involved in cortical maturation and migratory activity [1,41,42].

Furthermore, metabolomics, a post-genomic technique, has emerged as a valuable tool for assessing health, growth, and development, especially in the neonatal period [43,44]. Metabolomic profiling through the analysis of body fluids, such as urine, provides quantitative information about metabolic pathways and gene-environment interactions [45]. Urinary metabolomics holds promise for early identification of infants at risk for brain damage and understanding the pathogenesis of impaired brain development [46–51].

The combination of neuroimaging techniques, such as EEG, functional magnetic resonance imaging (fMRI) of the brain, along with metabolomics, offers a comprehensive evaluation approach in neonatology. These techniques provide real-time information on complex body systems, allowing for personalized diagnosis, therapy, and outcome prediction [51–54]. Metabolomics analysis complements the understanding of metabolic networks and facilitates clinical decision-making, enabling timely and individualized interventions [44].

This study aims to investigate the potential benefits of therapeutic and affective touch in promoting cortical development in preterm infants born between 32.0 and 33.6 weeks of gestation. Specifically, the objective is to determine whether repeated manual techniques, such as OMT, compared to affective touch and static touch, can induce and modify the brain connectome and its metabolism, as evidenced by biomarkers, in preterm newborns.

The hypothesis posits that both manual touch and affective touch can effectively improve neurophysiological parameters in premature newborns. Recent research suggests that OMT may interact with neurological structures by stimulating CT fibers, thereby modifying interoceptive afferents and their impact on various physiological functions, including the gastrointestinal region.

If the findings demonstrate that OMT and affective touch can indeed alter the quality of brain connections and metabolic biomarkers, it could provide valuable insights for developing strategies to enhance neurodevelopment and prevention in this vulnerable population. Preterm infants are more susceptible to neurodevelopmental issues compared to full-term infants, thus emphasizing the significance of exploring interventions that can positively impact their long-term outcomes.

## METHODS AND ANALYSIS

The current protocol is designed for a randomized sham-controlled pilot study enrolling premature babies born at the Buzzi Children’s Hospital in Milan in the period between January 2025 and March 2026 who meet the following inclusion criteria:

- Preterm birth, between 32.0 and 33.6 weeks gestational age (GA);
- Absence of comorbidities that could affect the stability of vital parameters, and therefore represent a contraindication to the proposed intervention. Comorbidities include sepsis, pathologies pertaining to surgery, respiratory or cardiovascular instability, birth from a drug-addicted or HIV-positive mother or known congenital pathologies;
- Obtaining informed consent for participation in this research project from parents or legal guardians.

Preterm infants born before 32.0 and after 34 weeks’ GA and with respiratory and neurological pathologies and any additional comorbidities will be excluded. Children whose parents will not read and sign or in case of failure to obtain informed consent will be excluded from the study. The protocol has been designed following the SPIRIT guidelines [55,56].

### Recruitment

The recruitment process for this study will involve enlisting premature infants from the Neonatology Department of Buzzi in Milan under the supervision of an expert neonatologist. The neonatologist will exercise discretion in determining the inclusion or exclusion of infants in the study based on predefined criteria. Once patients are enrolled in the study, they will receive standard medical treatment in accordance with established protocols. Additionally, the infants will undergo tactile stimulation as described in the study protocol, which includes specific techniques and procedures.

### Study design

The current trial employs a randomized, sham-controlled design to investigate the effects of touch interventions on preterm infants. The experimental group will be subjected to either therapeutic touch (OMT) or affective touch (AT), while the control group will receive non-specific static touch (No T).

The timeline of the study involves several time points (Figure 1). At T0, subjects will be recruited, and informed consent will be obtained from parents or legal guardians. At T1, urine samples will be collected from the infants before the interventions, whilst an EEG will be conducted during the interventions. Subsequently, from T2 to T4, urine samples will be collected before and after each touch session to compare pre-post intervention metabolomic outcomes. The touch intervention, therefore, will be administered three times at T2, T3, and T4, corresponding to 4, 8, and 12 days after T1, respectively.

**Figure 1.**
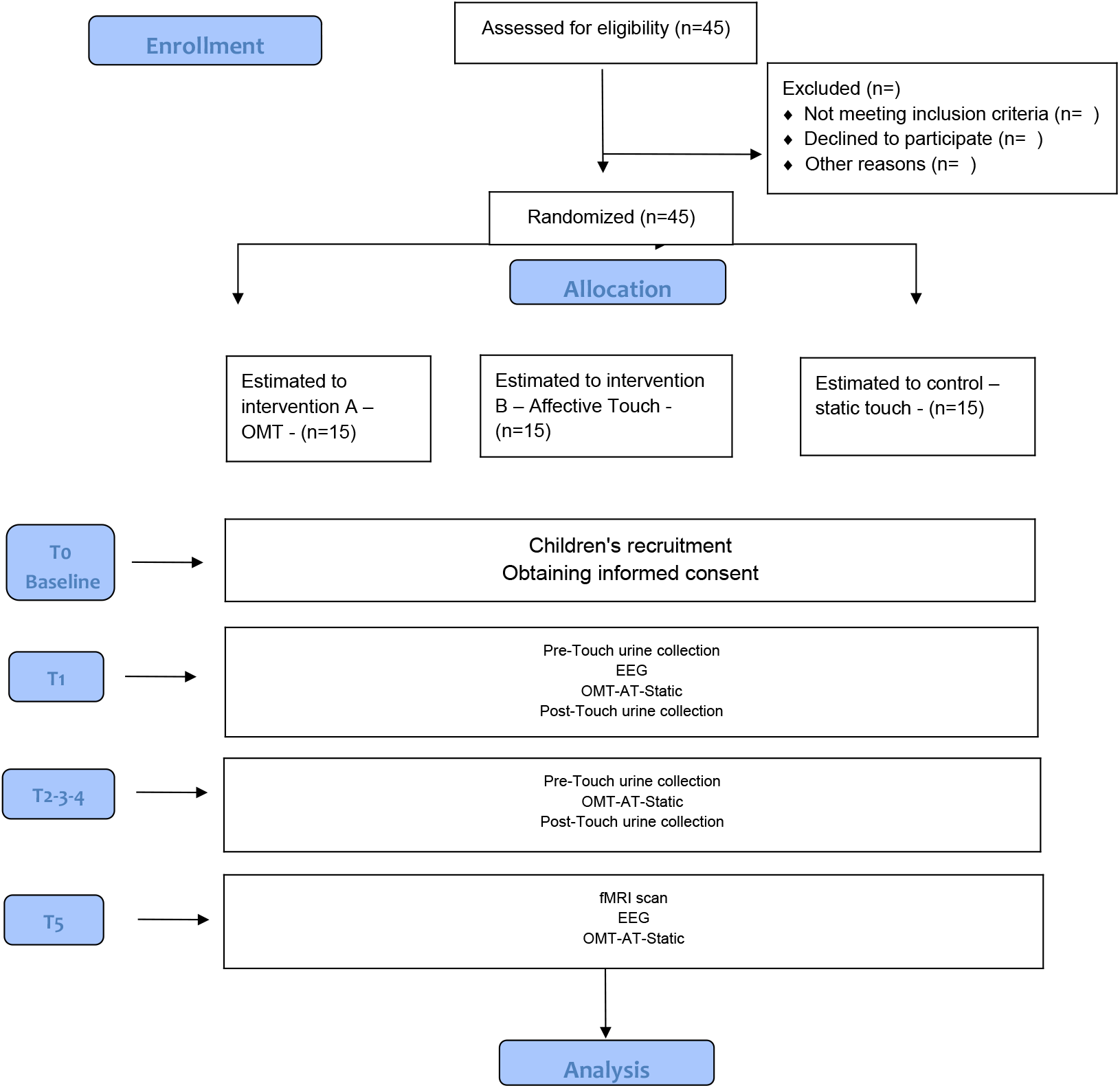
Flowchart of the study protocol. OMT=osteopathic manipulative treatment; AT= affective touch; EEG= Electroencephalogram.

At T5, corresponding to the 40th week of gestation, the infants undergo routine fMRI of the brain and EEG sessions. Additionally, during the fMRI and EEG procedure, the infants will receive the touch session based on the randomization they were assigned at T1. This will enable the examination of the immediate effects of different types of touch on brain activity.

### Randomization

All patients included in the study are registered with a progressive identifier (patient code). The treatment arm will be assigned through a random selection procedure. A computerized randomization algorithm based on the trinomial distribution builds a balanced and progressive list of random values assigned within a specific interval (in this case 2=OMT group; 1=affective touch group, 0=control/static touch group), also called randomization code. A progressive index is associated with each value. The treatment is assigned by attributing to each patient the randomization code relating to the equivalent patient code.

The randomization process will be based on the use of a randomization software (R statistical software) and carried out through a “permuted block” procedure with a 1:1:1 ratio.

The study includes three groups (one OMT study, one affective touch study and one static touch control) of 15 subjects each, for a total of 45 infants.

### Blinding/masking

Randomization will be carried out by the Neonatal Intensive Care Unit which will control all processes. Personnel blinding will be respected through the following principles: a single MD will carry out the initial and final evaluation of the subjects included in the study. Operators performing the brain fMRI, EEG, and metabolomics measurements will be unaware of the subjects in the study and control groups. None of the operators involved will be aware of the purpose and design of the study. The subjects included in the study will not be aware of the group they belong to given the inclusion criteria relating to treatment with manual therapy techniques. The blinding of the operators making the touch cannot be maintained given the active nature of the treatment.

### Outcomes

The primary outcome for the present protocol is the pre-post changes between groups in BOLD levels among different brain areas, specifically the anterior insula and the medial prefrontal cortex. BOLD connectivity will be evaluated among 90 subcortical and cortical ROIs defined by the University of North Carolina (UNC) Infant Atlas [57].

A series of secondary outcomes have been planned and included: pre-post changes in BOLD levels and connectivity within the salience network (SN) between the study and control groups; intragroup and intergroup differences in resting-state networks; pre-post changes from baseline in the EEG power in slow delta waves band at the end of the treatment period (T5-T1); pre-post changes in urinary metabolites as assessed by Proton Nuclear Magnetic Resonance Spectroscopy (1H NMR) at different timpoints, such as T1, T2, T3, T4, T5; pre-post changes in urinary metabolites as assessed by Mass Spectrometry combined with Liquid Chromatography (CL-MS) at T1, T2, T3, T4, T5; pre-post changes in urinary metabolites as assessed by Mass Spectrometry combined with Gas Chromatography (CG-MS) at T1, T2, T3, T4, T5 (Figure 2).

**Figure 2.**
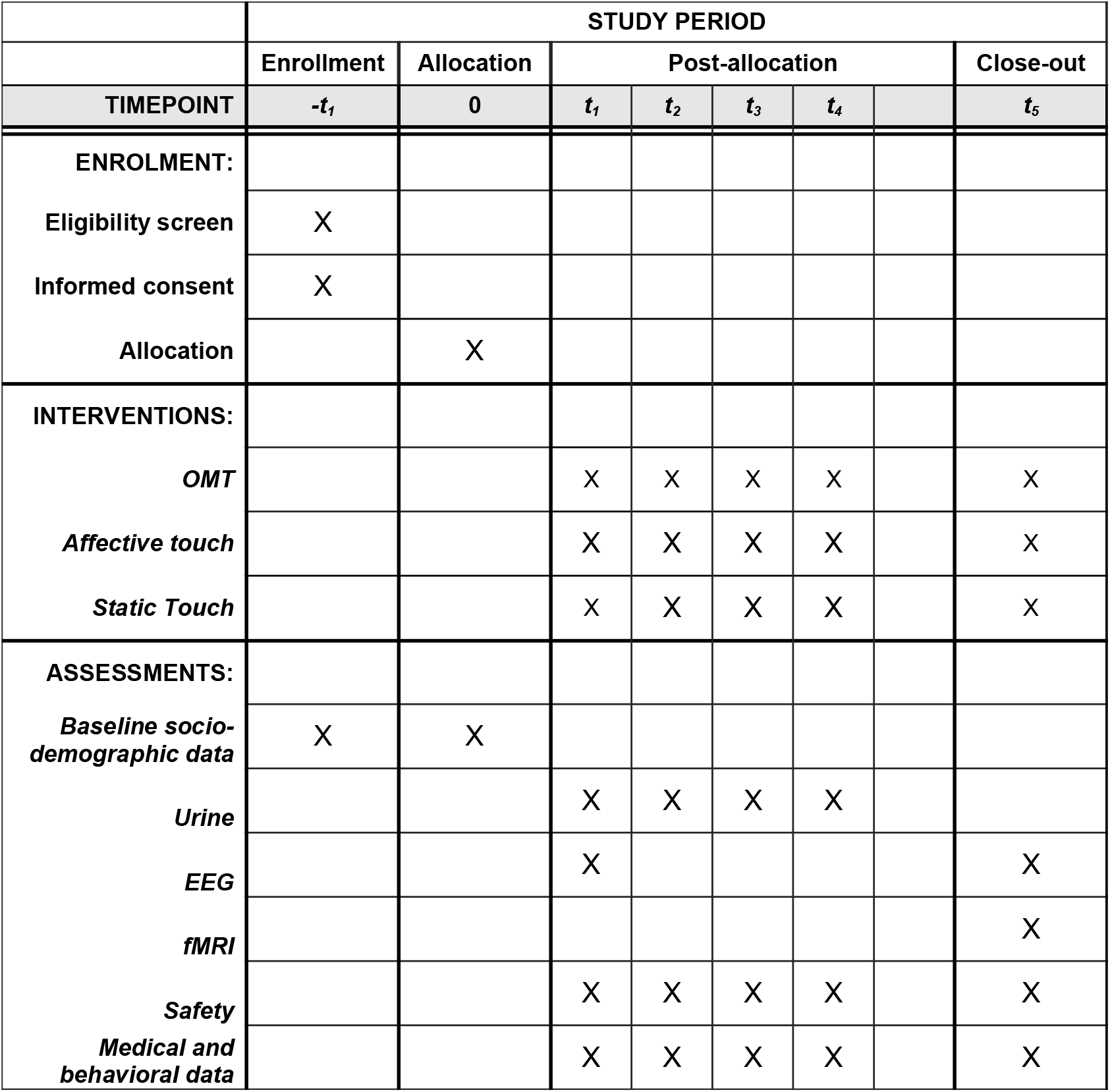
Template of recommended schedule of enrollment, interventions, and assessments. OMT=Osteopathic Manipulative Treatment; EEG=Electroencephalogram; fMRI: Functional magnetic resonance imaging

#### Metabolomics

Urinary metabolomics has the potential to serve as a noninvasive biomarker of brain metabolism.

The collection of urine will be tested pre and post each timepoint to determine eventual variation among brain chemicals, as a metabolic product, after the administration of the various types of touch, as outlined in Figure 1 and 2.

#### Safety

Monitoring adverse events as measured by abnormal heart rate and oxygen levels patterns at each treatment session will be used to evaluate safety (Figure 2).

### Data collection strategy

To mitigate potential confounding variables between hospital discharge and the T5 assessment, a structured parental diary and a standardized electronic data collection system will be implemented. Strategies will be employed to ensure reliability, minimize bias, and enhance adherence, including parental pre-training, automated reminders, and standardized reporting formats.

#### Data Collection Procedures

Data will be collected every other day via a structured anonymized in-house survey sent to participating families. To ensure data accuracy and reduce variability, parents will receive structured training before data collection begins, covering measurement techniques, reporting consistency, and the importance of minimizing recall bias. Every participant will be associated to an ID code to respect anonymicity and privacy according to the international guidelines and standards.

Automated weekly reminders and alerts will be sent to parents to enhance adherence and reduce missing data, in alignment with best practices for data collection.

#### Recorded Variables

The following variables will be systematically collected and categorized to facilitate statistical adjustment for potential confounders:

Anthropometric Measurements: Weekly documentation of infant weight, length, and head circumference, following standardized measurement protocols.

Daily Tactile Stimulation: Parents will report the duration of daily physical contact using predefined categories to reduce recall bias: <30 minutes; 30–60 minutes; 60–120 minutes; 120-180 minutes; >180 minutes.

Health Status: The occurrence of illnesses will be documented to assess disease burden.

Trauma Exposure: Parents will report any significant injuries, including falls, head trauma, and other adverse events, ensuring detailed classification for severity and impact.

Sleep-Wake Patterns: Sleep duration and wakefulness cycles will be recorded to assess potential disruptions or irregularities.

Data validation procedures, including plausibility checks and follow-up with parents for missing or inconsistent responses, will be implemented to enhance data integrity.

### Data acquisition

Magnetic resonance (MR) imaging data will be acquired using a 3 Tesla whole-body system (Siemens – Vida 3T) equipped with an 32-channel head-only receiver coil during natural sleep [58,59]. Neonates will be positioned supine within the scanner and securely wrapped in blankets, with molded foam utilized to restrict head movement. To safeguard against auditory stimulation, neonatal earmuffs and custom-fitted ear-canal plugs will be employed. Continuous monitoring of heart rate and oxygen saturation will be conducted by an attending phisician throughout the MR imaging session. Structural imaging data will be acquired utilizing a T1-weighted Turbo Field Echo (TFE) sagittal sequence (flip angle: 8°; TR: 9 ms; TE: 4.2 ms; voxel size: 1 × 1 × 1 mm^3; FOV: 200 × 200 × 150 mm^3), maintaining a whole-body specific absorption rate (SAR) below 0.2 W/kg. Following the completion of standard clinical MRI sequences, whole-brain functional images will be obtained using a T2*-weighted echo-planar imaging (EPI) sequence (flip angle: 90°; TR: 1555 ms; TE: 30 ms; voxel size: 2.5 × 2.5 × 3 mm^3; FOV: 180 × 180 × 75 mm^3; slice gap: 0 mm), while ensuring a whole-body SAR within 0.8 W/kg.

A measurement will be taken when the 40th week is reached. The sequences that will be used are: T2-weighted echo-planar* for fMRI measurements with Blood Oxygenation Level Dependent (BOLD) contrast and pseudo-continuous Arterial Spin Labeling (pCASL) for cerebral blood flow (CBF) measurements.

The sequences of the resting state will be obtained during the period of tactile stimulation (different for each subject’s group). The anatomical target of the fMRI analysis will be the anterior insula. Parents/legal guardians will fill out a specific safety questionnaire for the MR examination, as is standard practice according to the standard operating procedures adopted in the clinical practice of the Buzzi hospital.

The EEG examination will be conducted at T1 and T5 utilizing a 50 channel equipment with electrodes placed on the child’s head following international norms and guidelines. The first five minutes of baseline will be recorded without contact. After that, the manual techniques or static contact outlined in the “intervention” section will be applied. The EEG acquisition will continue during this time. Once finished, there will be additional 5 minutes post-touch recording. The total time for the EEG examination is estimated as approximately 1 hour.

Urine sample collection and storage procedure for metabolomic analysis

Urine samples will be collected before and after each intervention at every timepoint. The collecting procedure will be as follows:

- Urine collection will use a cotton ball, placed in the baby’s diaper, from which it is aspirated using a syringe (without a needle);
- Transfer of the urine collected (about 2 mL) into two 1.5 mL eppendolf tubes (1 mL per tube), in which sodium azide (NaN3) is already present.
- The decision to collect two samples per assessment is to allow each set to be analyzed with two methods: Nuclear Magnetic Resonance (NMR) and Mass Spectrometry associated with gaseous and/or liquid chromatography (GC/MS; LC /MS)
- The tubes will be then labeled, using indelible markers, with the identification of the subject and the collection time (T1, T2, T3, T4, T5), specifying whether the collection took place pre or post-intervention.
- Subsequently, the samples will be stored in the freezer at −80°C until shipment, which will be taking place with appropriate packaging in dry ice.

Analysis of metabolomic markers

The samples are analyzed by the Laboratory of the Humanitas University. The following methodologies will be using for analysis:

- Proton Nuclear Magnetic Resonance Spectroscopy (1H NMR),
- Mass Spectrometry combined with Liquid Chromatography (CL-MS),
- Mass Spectrometry combined with Gas Chromatography (CG-MS).

The choice to combine these techniques is given by the need to obtain the widest possible spectrum of information from the biological material taken [45,60].

The decision to collect samples prior to and after each intervention is dependent on the parameters of the analysis technique itself: metabolomics enables the detection of the metabolic state in real time and, consequently, facilitates the evaluation of its evolution over time.

### Procedure for data collection and processing

Data collection will be carried out using a specific software, created ad-hoc for the purpose of the study. The data will be treated according to the privacy law. All information will be encrypted, codified and anonymized. The acquired data will be used exclusively for the specific purpose of the research.

### Study arms

The preterm infants participating in the experimental arms of the study will be randomly assigned to receive either therapeutic touch or affective touch. The affective touch protocol is based on the principles of affective touch as described by McGlone (2014). A trained operator with a minimum of five years of experience in neonatology and at least eight hours of practical training in performing affective touch will gently touch the infant’s body using their right hand, maintaining a consistent speed of approximately 3 centimeters per second. The operator will wear a glove, and the temperature of the hand wearing the glove will be maintained between 32 and 36 degrees Celsius. The touch will be applied near the infant for a duration of 5 minutes, following the methodology described by Manzotti and colleagues [61–67].

During the scheduled instrumental examinations at T1 and T5, when brain fMRI and EEG tests are conducted, the newborn will be positioned accordingly, and the touch will be administered without altering the infant’s posture in areas of the body where contact is feasible, specifically focusing on the lumbar and pelvic regions based on previous studies [62,68]. The same procedure outlined above will be followed at all other time points.

For therapeutic touch, specifically osteopathic manipulative treatment, the approach will be adapted to the infant’s position during the session. A physiotherapist with a bachelor’s degree in health and specialized training in osteopathy, including at least two years of pediatric and neonatal manipulation training encompassing a minimum of 250 training hours, will administer the treatment. The osteopathic approach chosen is that already used in previous studies [19,69–71] and has proven to be safe in the context of preterm infants. Specifically, indirect techniques will be used (e.g. cranial ligament tension, functional, balanced).

While ensuring the child’s initial position is maintained, the operator may modify the contact surface of their hands as needed, taking care not to interfere with the designated protocol tests.

To standardize the duration of contact between the two types of touch, approximately 5 minutes will be allocated for their application.

In the control group, infants will receive static touch. The static touch will be administered to areas of the body that are accessible and free from procedural obstructions, following the same duration as the touch sessions in the study groups, which is 5 minutes. The operator will utilize the same methods employed for therapeutic and affective touch, ensuring that the fingers of the hand wearing the glove make contact with an unrestricted part of the child’s body and maintain that position throughout the 5-minute protocol.

### Statistical plan

#### Sample size

The absence of consolidated effect size estimates for manual treatment on the primary outcome presents a methodological challenge for precise sample size calculation. Despite increasing research interest, variability in study designs, outcome measures, and population characteristics complicate reliable parameter extrapolation for rigorous power analysis.

Given these limitations, we adopted a pragmatic approach based on prior literature, which suggests pilot studies typically include 10–20 subjects per group to provide preliminary effect size and variance estimates [72,73]. Accordingly, we selected 15 subjects per group, balancing feasibility with sufficient data collection to:

1. Assess primary outcome variability,
2. Estimate a preliminary effect size, and
3. Evaluate study logistics, including recruitment, adherence, and procedural feasibility.

This sample size aligns with previous pilot studies in similar contexts [74,75]. Given the exploratory nature of this study, results will serve as hypothesis-generating rather than conclusive, guiding the design of a larger, statistically powered clinical trial.

#### Statistical analysis

The statistical analysis will be based on the ‘intention-to-treat’ and ‘per-protocol’ methods. The statistical analysis will use the mean and standard deviation, median and percentiles, percentage points for the description of the variables of the three groups. Hypothesis H0 (null hypothesis) is no difference between groups at the end of treatment. To test this hypothesis we will first test the assumption of the normal distribution of the data using the Levene test.

The analysis of the residuals will indicate the possible use of logarithmic transformations of the data. Subsequently, the univariate analysis will involve the use of the analysis of variance (ANOVA) for the comparison of the numerical variables, while the chi square (chi squared test) for the categorical and/or ordinal variables. Primarily used for the comparison of the baseline samples, with the same method the data at the end of the treatment will also be analyzed taking into account the impossibility of revealing the independent effect of the variables on the outcome. ANCOVA will be used to explore in more detail the treatment effect considering cofactors. A linear mixed effect model will explore the effects across groups.

Data will be analyzed within and between the three study groups. Furthermore, a linear regression model will be applied to investigate the independent effect of OMT on primary and secondary endpoints between study groups.

The level of statistical significance is established for an alpha < 0.05. The calculation of the power will be performed post-hoc. R statistical program will be used for data analysis.

## ETHICAL AND DISSEMINATION

The parents of the infants who will participate in the research will be informed about the objectives and methods of conducting the study through a verbal interview and the signing of the informed consent.

Ethical approval has been obtained from the Ethics Committee of the local health agency in Milan (CET 449-2024). The research protocol has been registered on ClinicalTrials.gov (NCT05853991) as of May 2, 2023. Considering the recent osteopathic literature [76–78], no disadvantages or anticipated adverse events and side effects related to OMT are expected. However, throughout the research period, any observed side effects and reasons for participant dropouts will be diligently documented and discussed in the final publication.

To date, no previous osteopathic trial has been conducted in this area with the intention of quantifying the benefits of OMT on newborns’ brain functions and metabolomics using rigorous procedures and gold-standard methods for clinical trials. This trial represents the first concrete experimental investigation addressing this outcome. The anticipated advantages of this trial include employing diverse methods to accurately quantify the physiological effects of OMT compared to other forms of touch, opening new avenues for investigating metabolic effects in moderately preterm infants in good clinical condition and without severe comorbidities, and contributing novel insights to the field of osteopathy research in newborns.

Dissemination of our study findings will be conducted through a comprehensive strategy targeting various stakeholders, including healthcare professionals, researchers, policy makers, and patient communities.

Primary dissemination channels will involve publications in peer-reviewed journals and presentations at national and international conferences, ensuring the scientific community is informed of our results.

Recognizing the ethical imperative to share outcomes with study participants, we will provide accessible summaries of our findings to the parents or legal guardians of participating infants, adhering to best practices in participant communication. Additionally, we will engage with patient advocacy groups and utilize digital platforms to broaden the reach of our findings, facilitating the translation of research into clinical practice and informing future guidelines for neonatal care.

## Data Availability

All data produced in the present study are available upon reasonable request to the authors

## Acknowledgment

The authors thank Drs. Paola Sciomachen, Mauro Longobardi and Marco Petracca for the invaluable help in supporting the project.

## Trial status

The present protocol is for an ongoing research.

## Author Contributions

The study was conceptualized and designed by AM, FC, and GL, with DG, PV, VF, AR, and FA contributing to the refinement of study questionnaires and overall design. Data collection will be carried out by EL, AR, FA, VF, MR, and PV. Statistical analyses will be performed by FC, FA, MR, and PV, in collaboration with professional statisticians. The initial draft of the manuscript was prepared by AM and FC, while all authors provided critical revisions and approved the final submitted version.

